# Respiratory Syncytial Virus (RSV) RNA in wastewater settled solids reflects RSV clinical positivity rates

**DOI:** 10.1101/2021.12.01.21267014

**Authors:** Bridgette Hughes, Dorothea Duong, Bradley J. White, Krista R. Wigginton, Elana M. G. Chan, Marlene K. Wolfe, Alexandria B. Boehm

## Abstract

Wastewater based epidemiology (WBE) uses concentrations of infectious agent targets in wastewater to infer infection trends in the contributing community. To date, WBE has been used to gain insight into infection trends of gastrointestinal diseases, but its application to respiratory diseases has been limited to COVID-19. Here we report Respiratory Syncytial Virus (RSV) genomic RNA can be detected in wastewater settled solids at two publicly owned treatment works (POTWs). We further show that its concentration in settled solids is strongly associated with clinical positivity rates for RSV at sentinel laboratories across the state in 2021, a year with anomalous seasonal trends in RSV disease. Given that RSV infections have similar clinical presentations to COVID-19, can be life threatening for some, and immunoprophylaxis distribution for vulnerable people is based on outbreak identification, WBE represents an important tool to augment current RSV surveillance and public health response efforts.

**Graphical Abstract:** 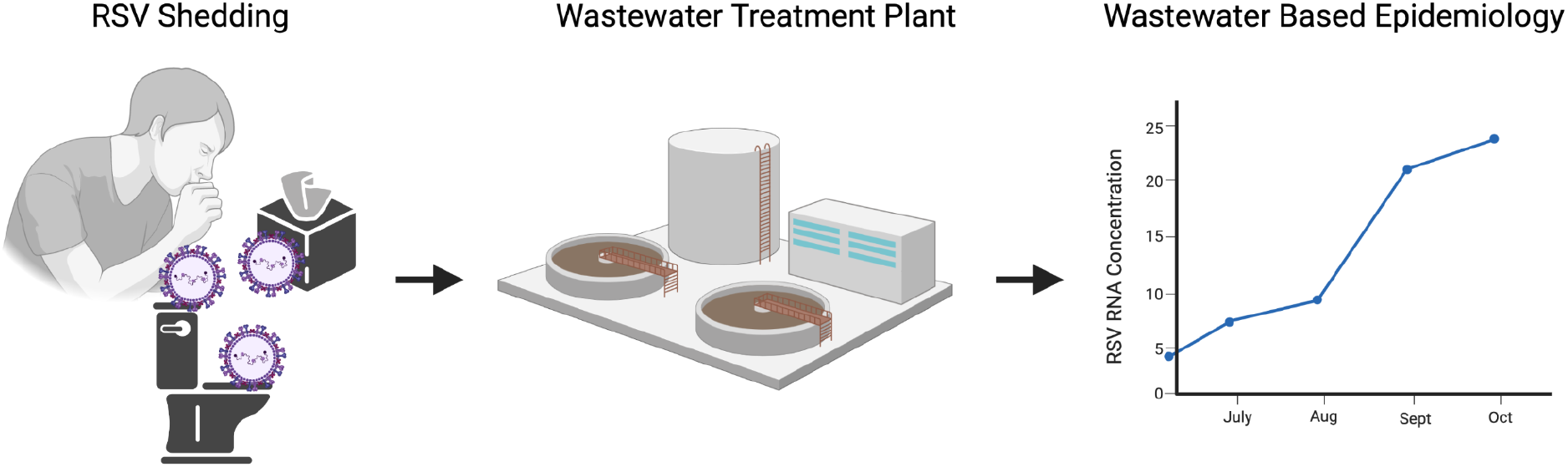

**Synopsis:** Respiratory Syncytial Virus RNA concentrations in settled solids from wastewater treatment plants are associated with state-wide RSV clinical positivity rates.

## Introduction

Respiratory syncytial virus (RSV) is a single-stranded, - sense RNA, enveloped virus that infects the respiratory tract of humans. RSV infections are usually self-limiting in healthy individuals but can become severe, particularly for infants and immunocompromised or elderly individuals. Symptoms include congestion, dry cough, fever, and trouble breathing in severe cases. Up to 1-2% of infants under 6 months with RSV are hospitalized, and RSV is responsible for up to a third of hospitalizations for acute respiratory infections (ARI) in children <5 in the United States^1^. RSV is reportable to public health officials in some locations in the United States, but many RSV infections are not identified because people do not require or seek care and testing is not always done when patients encounter the healthcare system. Given the similarities in presentation between COVID-19 and RSV, the challenge in obtaining data on RSV in communities, and the high risk associated with infection in infants, there is a need to better understand RSV incidence among US populations.

Wastewater-based epidemiology (WBE) is a population health monitoring approach that uses concentrations of pathogenic agents in wastewater to gain insight into infection rates in the contributing community^2^. Using wastewater to understand community infection rates is a passive method that has the benefit of capturing inputs from an entire community without a participation burden. WBE has been used to gain insight into community infection rates of non-enveloped viruses including poliovirus^3^, norovirus^4^, and hepatitis A^5^, which are excreted in stool of infected individuals and transmitted via the fecal-oral route. Widespread implementation of WBE since the beginning of the COVID-19 pandemic shows strong associations between SARS-CoV-2 RNA in wastewater influent^6–8^ and settled solids ^9–11^ and laboratory-confirmed COVID-19 incidence rates. Although COVID-19 is primarily a respiratory disease, SARS-CoV-2 RNA is excreted in stool of those infected^12–14^. To date WBE has not been used to provide information on infection rates of other respiratory viral diseases such as RSV.

Population monitoring for RSV can help establish guidelines for diagnostic testing and preventative measures. RSV is a seasonal outbreak disease with infections typically peaking in October and transmission ongoing through May, however seasonality can differ across regions of the United States and from year to year^15^. The 2020/2021 RSV season in the United States was substantially delayed during the COVID-19 pandemic and in June 2021 the US Centers for Disease Control and Prevention issued a health advisory warning of unexpected interseasonal RSV outbreaks^16^. Limited RSV immunoprophylaxis products are currently available to those at high risk and administered on a monthly basis depending on both personal risk and outbreak status^17^. Identifying outbreaks is thus important for targeting administration of these monoclonal antibodies, and for ongoing vaccine trials for multiple promising candidates and eventually targeting vaccine administration.

To investigate whether RSV is present in stool of infected patients and thus potentially present in wastewater, we conducted a literature review (see Supporting Information (SI)). We identified two peer-reviewed papers that tested for the presence of RSV genomic RNA in stool samples. von Linstow et al.^18^ studied 44 children admitted to a hospital over a 6 month period with an ARI caused by RSV or human metapneumovirus. They collected one stool sample at enrollment and then weekly for three weeks from each child. RSV viral RNA was detected using quantitative RT-PCR in 5 of 165 total stool samples. The five positive stool samples were obtained from five unique children with RSV infections. Shedding in stool only occurred within the first week of illness. In 2017, a case report by Akbari et al.^19^ detected RSV RNA in stool. Three stool samples were collected from a 12-month old female presenting with acute gastroenteritis and tested for respiratory viruses and gastrointestinal bacteria using a complete multiplex-PCR panel. RSV (subtype B) was present in all three stool samples. Neither study reported RSV RNA concentrations in stool, or provided detection limits for their assays.

The goal of this study is to investigate whether RSV can be detected in wastewater and, if so, whether its concentration is associated with clinical RSV positivity rates. Previous work indicates that enveloped viruses partition to the solids in wastewater where their concentrations are enriched orders of magnitude over the liquid phase^20,21^. We therefore focused our analysis on wastewater solids at two publicly owned treatment works (POTWs) in a highly urbanized region of the San Francisco Bay Area, California, USA. We first designed and tested a custom digital droplet (dd-)RT-PCR assay that targets the N gene of RSV A and RSV B. We then applied the assay to settled solids collected biweekly to three days per week at the POTWs between January and mid-November 2021. The resultant concentrations are compared to the limited information publicly available on RSV infections in the State of California.

## Methods

### Assay development for RSV

A digital RT-PCR assay was designed to target the N gene of RSV. RSV A and RSV B genomes were downloaded from NCBI. The assay was developed *in silico* using Primer3Plus (https://primer3plus.com/). The parameters used in assay development (that controlled sequence length, GC content, and melt temperatures) are provided in Table S1.

Primers and probe sequences were screened for specificity in silico using NCBI Blast, and then tested in vitro against a virus panel (NATtrol™ Respiratory Verification Panel NATRVP2-BIO, Zeptomatrix, Buffalo, NY) that includes several chemically inactivated intact influenza viruses, parainfluenza viruses, adenovirus, rhinovirus, metapneumovirus and several coronaviruses, and “wild-type” gRNA from SARS-CoV-2 strain 2019-nCoV/USA-WA1/2020 (ATCC® VR-1986D™). A mixture of RSV B (NATtrol™ RSV positive Control NATRSV-6C, Zeptometrix) and RSV A (from NATtrol™ Flu Verification Panel NATFVP-NNS, Zeptometrix) was also included as a positive control. RNA was extracted using Chemagic™ Viral DNA/RNA 300 Kit H96 for Chemagic 360 (Perkin Elmer, Waltham, MA). RNA was used undiluted as template in digital droplet PCR singleton assays. The concentration of targets used in the in vitro specificity testing was between 10^3^ and 10^4^ copies per well; each target from the respiratory panel was run in a single well, and the SARS-CoV-2 gRNA was tested in 8 wells.

### Wastewater samples

Two publicly owned treatment works (POTWs) that serve populations of Santa Clara County, California, USA (San José-Santa Clara Regional Wastewater Facility, RWF) and San Mateo County, California, USA (Silicon Valley Clean Water, SVCW) were included in the study. RWF and SVCW serve approximately 1,500,000 and 220,000 people, respectively; further descriptions of these POTWs can be found in Wolfe et al.^22^.

Samples of approximately 50 mL of settled solids were collected by POTW staff using sterile technique in clean, labeled bottles. Grab samples were taken at SVCW. At RWF, POTW staff manually collected a 24 h composite sample^21^. Samples were immediately stored at 4°C, transported to the lab, and processed within 6 hours of collection.

Samples were collected daily for a larger COVID-19 wastewater surveillance effort starting in November 2020^22^, and a subset of these samples are used in the present study and were chosen to span the period prior to and including increasing RSV incidence in the state^23^. Although RSV incidence usually peaks in the winter, the 2020-2021 was an anomalous year with limited to no RSV cases and increasing cases beginning in the summer (Figure 1). One sample was analyzed every other week during January, February, and March; one sample was analyzed per week in April and May, and then three samples per week were analyzed between 22 May to 12 November 2021. 90 samples from each POTW were included in the analysis for a total of 180 samples.

**Figure 1.**
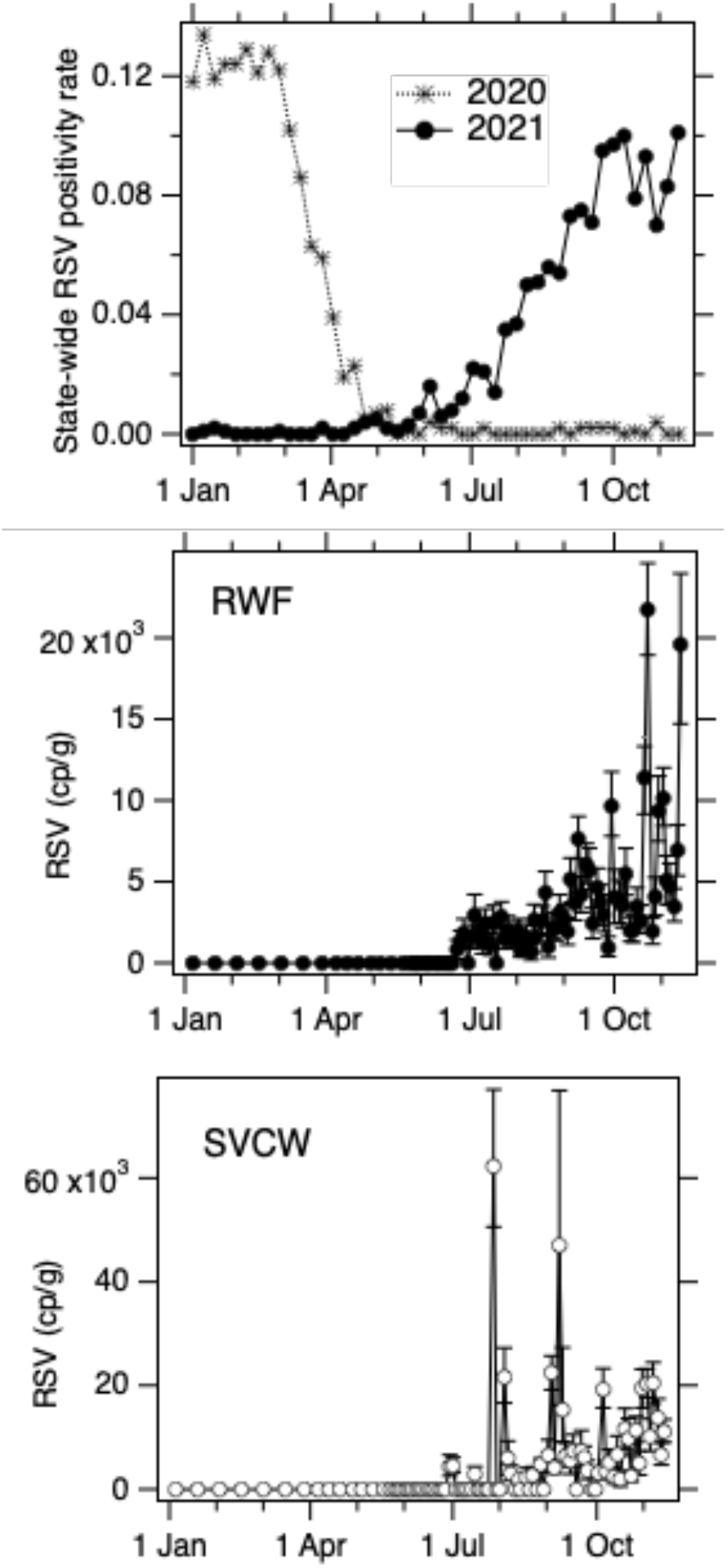
Concentrations of RSV RNA (units of copies per g dry weight) measured in settled solids at RWF and SVCW POTWs in 2021 as well as the percent of clinical samples from sentinel laboratories testing positive for RSV across the state of California in 2020 and 2021 (positivity rate). Error bars on the wastewater measurements represent standard deviations as total errors from the ddPCR instrument software.

RNA was extracted from the 10 replicate aliquots of dewatered settled solids as described in Wolfe et al. ^22,24,25^. RNA was subsequently processed immediately (within 24h of sample collection) to measure concentrations of the N gene of SARS-CoV-2, pepper mild mottle virus (PMMoV), and bovine coronavirus (BCoV) recovery using digital droplet RT-PCR methods reported by Wolfe et al. ^22,26^. PMMoV is highly abundant in human stool and wastewater globally^27,28^ and is used here as an internal recovery and fecal strength control for the wastewater samples^29^. BCoV was spiked into the samples and used as an additional recovery control; all samples were required to have greater than 10% BCoV recovery.

During the initial measurement, RNA extraction and PCR negative and positive controls were included to ensure no contamination as described in Wolfe et al.^22^. Extracted RNA samples were then stored at - 80°C between 4 and 300 days before they were analyzed for the RSV N gene and SARS-CoV-2 N gene in a multiplex digital RT-PCR assay; each of the 10 replicate RNA extracted were run in its own well, and the 10 wells were merged for analysis. The SARS-CoV-2 N gene was run a second time in all samples to test for RNA degradation during storage (no to minimal degradation was observed, see SI). Wastewater data and MIQE reporting details^30^ are available publicly at the Stanford Digital Repository (https://purl.stanford.edu/ck044th0182); results below are reported as suggested in the EMMI guidelines^31^.

### ddRT-PCR

For the assay specificity testing, the RSV assay was challenged against target (RSV A and B) and non-target (other respiratory viruses) viral gRNA. Each PCR plate contained positive controls and negative PCR no template controls (NTC). PCR positive controls for each target assayed on the plate were run in 8 wells, and NTC were run in 8 wells. Positive controls for the assays consisted of a mixture of RSV A and B gRNA.

The ddRT-PCR methods applied to wastewater solids to measure the SARS-CoV-2 N gene, PMMoV, and BCoV are provided in detail elsewhere, including details relating to EMMI^31^ guidelines and are not repeated here^22^. Here, we provide methods for the multiplexed SARS-CoV-2 N and RSV N gene assays which have not been reported previously. For this multiplexed assay, ddRT-PCR was performed on 20 μl samples from a 22 μl reaction volume, prepared using 5.5 μl template, mixed with 5.5 μl of One-Step RT-ddPCR Advanced Kit for Probes (Bio-Rad 1863021), 2.2 μl Reverse Transcriptase, 1.1 μl DTT and primers and probes at a final concentration of 900 nM and 250 nM respectively. Primer and probes for the RSV A & B N gene were purchased from Integrated DNA Technologies (IDT, San Diego, CA) (Forward primer: CTCCAGAATAYAGGCATGAYTCTCC, Reverse Primer: GCYCTYCTAATYACWGCTGTAAGAC, Probe: TAACCAAATTAGCAGCAGGAGATAGATCAG (5’ HEX/ZEN/3’ IBFQ)).

Droplets were generated using the AutoDG Automated Droplet Generator (Bio-Rad, Hercules, CA). PCR was performed using Mastercycler Pro (Eppendforf, Enfield, CT) with with the following cycling conditions: reverse transcription at 50°C for 60 minutes, enzyme activation at 95°C for 5 minutes, 40 cycles of denaturation at 95°C for 30 seconds and annealing and extension at 61°C for 30 seconds, enzyme deactivation at 98°C for 10 minutes then an indefinite hold at 4°C. The ramp rate for temperature changes were set to 2°C/second and the final hold at 4°C was performed for a minimum of 30 minutes to allow the droplets to stabilize. Droplets were analyzed using the QX200 Droplet Reader (Bio-Rad). A well had to have over 10,000 droplets for inclusion in the analysis. All liquid transfers were performed using the Agilent Bravo (Agilent Technologies, Santa Clara, CA).

Thresholding was done using QuantaSoft™ Analysis Pro Software (Bio-Rad, version 1.0.596). In order for a sample to be recorded as positive, it had to have at least 3 positive droplets. Each wastewater sample was run in 10 replicate wells, and each 96-well PCR plate of wastewater samples included PCR positive controls for each target assayed on the plate in 1 well, PCR NTCs in two wells, extraction negative controls (where no sample was added to the extraction kit) in two wells, and one extraction positive control (containing SARS-CoV-2 gRNA) in one well. PCR positive controls consisted of gRNA of SARS-CoV-2 (strain 2019-nCoV/USA-WA1/2020, ATCC® VR-1986D™) and RSV B and A gRNA (NATRSV-6C and RSV A from NATtrol™ Flu Verification Panel, Zeptometrix).

Results from replicate wells were merged for analysis. Thresholding was done using QuantaSoft™ Analysis Pro Software (Bio-Rad, version 1.0.596). In order for a sample to be recorded as positive, it had to have at least 3 positive droplets. For the wastewater samples, three positive droplets corresponds to a concentration between ∼500-1000 cp/g; the range in values is a result of the range in the equivalent mass of dry solids added to the wells.

For the wastewater samples, concentrations of RNA targets were converted to concentrations per dry weight of solids in units of copies/g dry weight using dimensional analysis. The dry weight of the dewatered solids was determined by drying^25^. The total error is reported as standard deviations and includes the errors associated with the Poisson distribution and the variability among the 10 replicates. A

### Clinical data in RSV

The weekly fraction of samples positive for RSV (hereafter “RSV positivity rate”) from sentinel laboratories in the State of California are publicly available from the California Department of Public Health website (https://www.cdph.ca.gov/Programs/CID/DCDC/Pages/Immunization/Influenza.aspx)^23^. Sentinel laboratories are hospital, academic, and private laboratories, and public health laboratories in the Respiratory Laboratory Network situated throughout California.

### Statistical analysis

RSV N gene concentrations were compared between POTWs using Kendall’s tau and Wilcoxon Sign Tests as data were not normally distributed (Shapiro-Wilk normality test, p<0.05). Weekly average RSV N gene concentrations and weekly average RSV N gene concentrations normalized by PMMoV were calculated for each week of 2021 through 12 Nov 2021 to match the frequency of RSV positivity rate reporting; the weekly average was calculated using all available data (between 1 and 3 data points). We tested the null hypothesis that weekly average wastewater concentrations were not associated with weekly positivity rates from sentinel laboratories using Kendall’s tau. We used linear regression to determine the rate of change of RSV positivity with RSV wastewater concentrations.

Measurements below the detection limit were replaced with 500 copies/g (the approximate detection limit). All statistical analyses were implemented in R studio (version 1.4.1106).

## Results and Discussion

### RSV Assay specificity

In silico analysis indicated no cross reactivity between the RSV assay and deposited sequences in NCBI. When challenged against the respiratory virus panel and SARS-CoV-2 gRNA no cross reactivity was observed; and the assay amplified gRNA from both RSV A and RSV B. Positive controls and NTCs run on the sample plate were positive and negative. These results suggest that the RSV ddRT-PCR assay is specific to both RSV subtypes.

### RSV RNA concentrations in wastewater solids

All positive and negative controls were positive and negative respectively, indicating assays performed well and without contamination. BCoV recoveries were higher than 10% and PMMoV concentrations within the expected range for the POTW suggesting an efficient and acceptable recovery of RNA during RNA extraction (Figure S1).

Concentrations of the RSV N gene in wastewater solids ranged from non-detect to 6.2×10^4^ copies/g dry weight. At both POTWs RSV levels were non-detect until late June at which time concentrations began to increase through the last sample collected in mid-November (Figure 1). Concentrations of RSV RNA were positively and significantly correlated at the two POTW (Kendall’s tau = 0.52, p<10^−12^). Paired, log_10_- transformed concentrations were not different between POTWs (paired, Wilcoxon signed rank test V = 988, p=0.9385, N = 90). Given the close proximity of the POTWs (located 40 km from each other), the similar concentrations may suggest similar RSV infection rates in their sewersheds.

The temporal trend in RSV RNA concentrations in settled solids mirrors state-wide trends in RSV positivity rate, identifying an infection pattern that differs from usual seasonal trends. Weekly averaged RSV RNA concentrations at each POTW were strongly correlated with the weekly positivity rate (tau = 0.77, p<10^−10^ for RWF, tau = 0.65, p<10^−7^ for SVCW, N=39). We repeated the analysis normalizing RSV RNA concentrations by PMMoV and found similar results. Normalizing by PMMoV can serve to correct for sample-to-sample changes in RNA extraction efficiency, concentrations during storage, or differences in the fecal strength of the wastewater ^22,32^ (Figure S2). A 1 log_10_ increase in RSV RNA is associated with an 0.08 (RWF) and 0.05 (SVCW) increase in RSV positivity rate (r^2^ = 0.89, p <10^−15^ for RWF; r^2^ = 0.70, p <10^−10^ for SVCW, N = 39) (Figure 2). The results are unchanged if the RSV concentrations are normalized by PMMoV.

**Figure 2.**
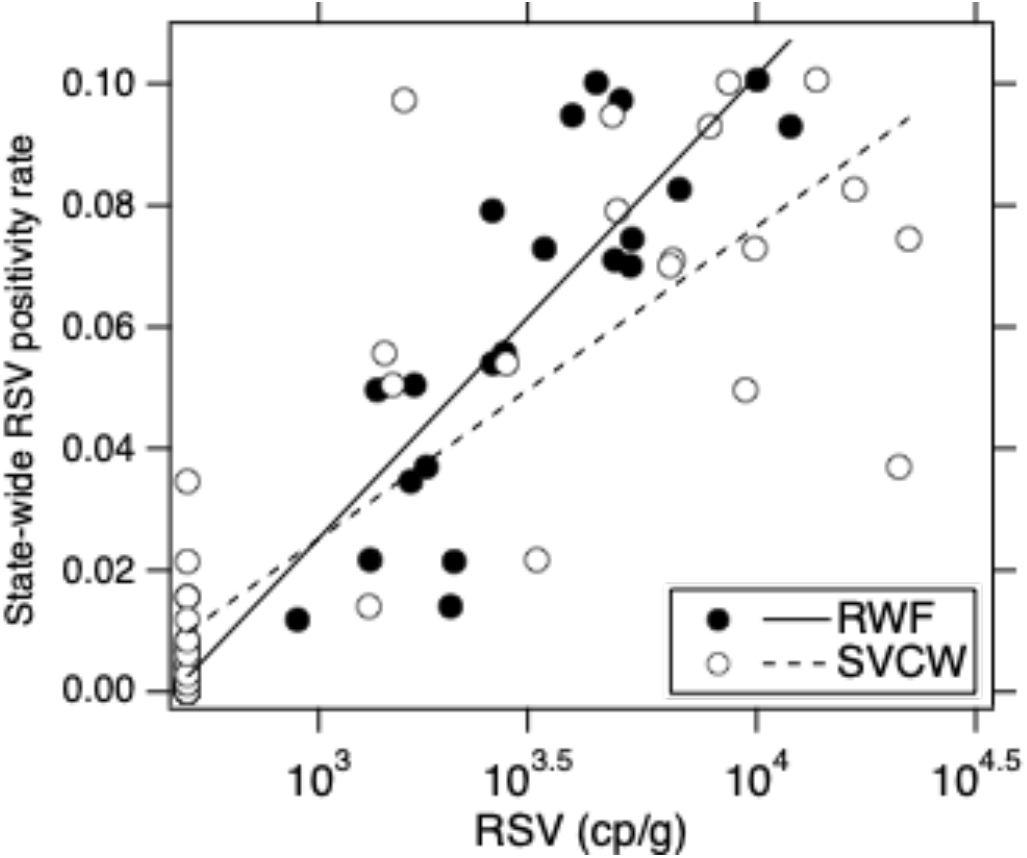
Weekly state-wide RSV positivity rate at sentinel laboratories versus weekly average RSV concentrations in wastewater settled solids at two POTW. Linear curve fit between log_10_-transformed concentrations and positivity rate is shown for both POTWs. Samples with RSV that are non-detect are shown with a value of 500 cp/g.

Concentrations of RSV RNA in wastewater solids reported are similar to concentrations of SARS-CoV-2 RNA in wastewater solids when laboratory-confirmed COVID-19 incidence rates are between 1/100,000 and 10/100,000^22^. The positivity rates used herein were calculated using data from sentinel laboratories through the state rather than from the sewershed areas, and thus do not provide direct information on population incidence rates. Additional work to compile RSV incidence data in the sewersheds may allow for estimates of disease burden associated with RSV in wastewater beyond outbreak identification and trends. Understanding how RSV RNA in wastewater relates to incidence rates will also require a better understanding of RSV shedding from infected individuals. In particular, there is no quantitative data on RSV RNA concentrations in stool of infected patients and the potential for sputum or other bodily secretions to contribute RSV RNA to wastewater should be considered.

The consistent detection of RSV in wastewater beginning during a known outbreak and strong associations between RSV RNA concentrations at the POTWs and state-wide trends in positivity rates at sentinel laboratories supports the idea that RSV RNA concentrations can be used to identify outbreaks and infer infection trends within contributing populations. Wastewater surveillance may be especially useful for diseases like RSV where clinical surveillance is challenging, yet tracking of outbreaks is required to initiate public health response. Wastewater concentrations may allow insight into when infection rates begin to rise, and may be especially useful for identifying unusual season trends in diseases like that observed for RSV in 2021. This work suggests that regular monitoring for RSV in wastewater can be useful to identify and target response to outbreaks. Success in monitoring for both COVID-19 and RSV suggests that additional efforts should be made towards wastewater monitoring for existing and novel enveloped respiratory viruses.

## Data Availability

All data are available at the Stanford Digital Repository as explained in the manuscript text.

## Acknowledgements

This work was funded by a gift from the CDC Foundation. We acknowledge the following individuals for assistance with wastewater solids collection: Payal Sarkar (RWF), Noel Enoki (RWF), and Amy Wong (RWF), Maria Gawat (SVCW), Tiffany Ishaya (SVCW), and Eric Hansen (SVCW). Graphical abstract Created with BioRender.com.

## Supporting Information

Additional methods, Tables S1 and S2 show primer design criteria and systematic review process, and Figures S1 and S2 show PMMoV results and RSV normalized by PMMoV, respectively.

