## Supporting Information for "Respiratory Syncytial Virus (RSV) RNA in wastewater settled solids reflects RSV clinical positivity rates"

Number of pages: 5
Number of figures: 2
Number of tables: 2

**Systematic review details.** We searched PubMed, Web of Science, and Scopus, and Google Scholar on October 13, 2021 for papers mentioning RSV and stool or feces (Table S2). Among the results, two papers were identified that tested stool for RSV RNA. Those two papers are described in the main text,

**Additional methods.**
The concentration of the SARS-CoV-2 N gene was measured immediately upon sample collection for a COVID-19 surveillance effort[1]. It was measured again in multiplex with the assay targeting the RSV N gene after RNA samples were stored for up to 11 months to gain insight into potential RNA degradation during storage. The concentration of SARS-CoV-2 RNA measured after storage at -80°C was not different from the concentration measured immediately after sample collection for the majority of the samples (96 of the 180 samples). For samples in which the concentration was different, the median ratio of the measurements was 0.78 suggesting minimal RNA degradation during storage.

Table S1. Parameters used with primer design software.

- Product size ranges: 60-200
- Primer size: min 15, opt 20, max 36
- Primer melting temperature: min 50°C, optimal 60°C, max 65°C
- GC% content: min 40%, optimal 50%, high 60%
- concentration of divalent cations = 3.8 mM
- concentration of dNTPs needs to be 0.8 mM
- Internal Oligo: size min 15, optimal 20, max 30
- Internal Oligo: Melting temp min 62°C, optimal 63°C, max 70°C
- Internal Oligo: GC% min 30%, optimal 50%, max 80%

Table S2. Search engines and search terms used in literature review.

| Search Engine | Search terms | Number of papers returned |
|---|---|---|
| PubMed | RSV AND (stool OR feces OR faeces) | 38 |
| Web of Science | (ALL=(RSV)) AND ALL=((stool OR feces OR faeces)) | 32 |
| Scopus | TITLE-ABS-KEY(RSV AND (stool OR feces OR faeces)) | 45 |
| Google Scholar | allintitle: stool OR feces OR faeces "respiratory syncytial virus" | 1 |

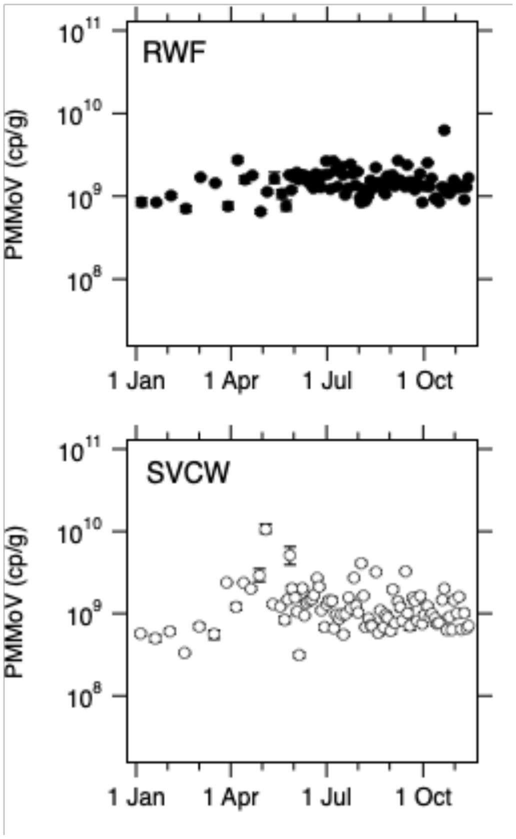

Figure S1. PMMoV concentrations in wastewater solids in units of copies per gram dry weight. Error bars represent standard deviations as total errors from the ddPCR instrument software. Some error bars cannot be seen because they are smaller than the symbol.

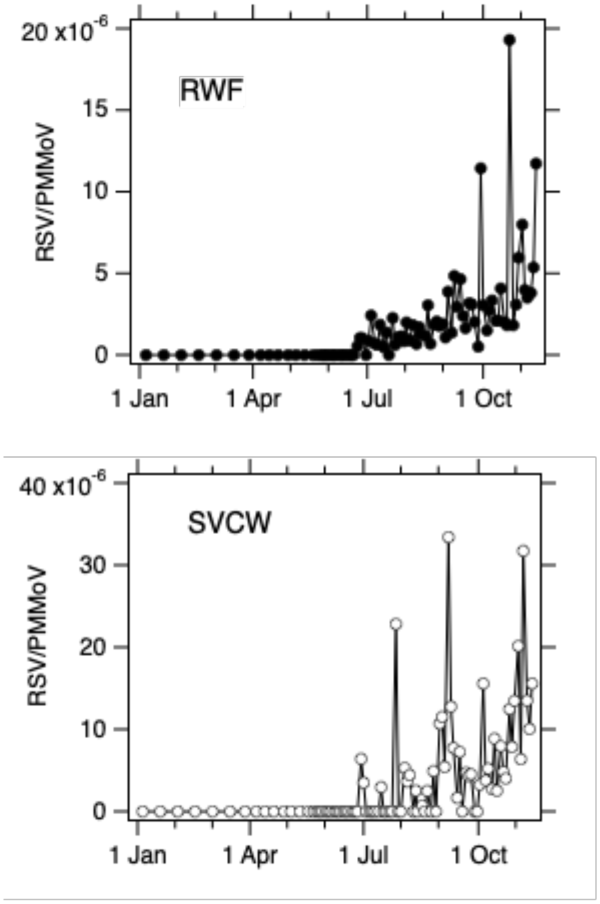


Figure S2. RSV normalized by PMMoV in wastewater solids.
